# Leveraging serology testing to identify children at risk for post-acute sequelae of SARS-CoV-2 infection: An EHR-based cohort study from the RECOVER program

**DOI:** 10.1101/2022.06.20.22276645

**Authors:** Asuncion Mejias, Julia Schuchard, Suchitra Rao, Tellen D. Bennett, Ravi Jhaveri, Deepika Thacker, L. Charles Bailey, Dimitri A. Christakis, Nathan M. Pajor, Hanieh Razzaghi, Christopher B. Forrest, Grace M. Lee, the RECOVER consortium

**Affiliations:** Division of Infectious Diseases, Department of Pediatrics, Nationwide Children’s Hospital and The Ohio State University, Columbus, OH; Applied Clinical Research Center, Children’s Hospital of Philadelphia, Philadelphia, PA; Department of Pediatrics, University of Colorado School of Medicine and Children’s Hospital Colorado, Aurora, CO; Division of Infectious Diseases, Ann & Robert H. Lurie Children’s Hospital of Chicago, Chicago, IL; Division of Cardiology, Nemours Children’s Health, Wilmington, DE; Center for Child Health, Behavior and Development, Seattle Children’s Hospital, Seattle, WA; Division of Pulmonary Medicine, Cincinnati Children’s Hospital Medical Center and University of Cincinnati College of Medicine, Cincinnati, OH; Department of Pediatrics (Infectious Diseases), Stanford University School of Medicine, Stanford, CA

**Keywords:** PEDSnet, COVID-19 serology, anti-N antibodies, anti-S antibodies, post-acute sequelae COVID-19, long COVID, Chronic COVID-19 Syndrome, Late sequelae of COVID-19, Long haul COVID-19, Long-term COVID-19, Post COVID-19 syndrome, Post-acute COVID-19, Post-acute sequelae of SARS-CoV-2 infection

## Abstract

The impact of post-acute sequelae of SARS-CoV-2 infection (PASC) in children is underrecognized. We developed an EHR-based algorithm across eight pediatric institutions to identify children with COVID-19 based on serology testing from 3/2020 through 4/2022 who had not been identified by PCR. Overall, serology tests were used 100-fold less than PCR. Seroprevalence of IgG anti-nucleocapsid antibodies remained stable, while rates of positive IgG anti-spike antibodies increased in teenagers after COVID-19 vaccine approval. Through data harmonization and after excluding 1,410 serology test results that may have been influenced by vaccines, we identified 2,714 children that were COVID-19 positive exclusively by serology. These patients were frequently tested as inpatients (24% vs. 2%), had chronic conditions more frequently (37% vs 24%), and a MIS-C diagnosis (23% vs. <1%) compared with PCR-positive children. Identification of children that could have been paucisymptomatic, not tested, or missed is critical to define the burden of PASC in children.

## INTRODUCTION

Since the beginning of the COVID-19 pandemic, different testing modalities for diagnosis have been implemented.^1^ While molecular tests remain the more reliable and accurate modality to diagnose acute SARS-CoV-2 infection, serology testing has key advantages. One, in the absence of a prior positive molecular test, it helps with the diagnosis of conditions that occur after SARS-CoV-2 infection, such as the multisystem inflammatory syndrome in children (MIS-C)^2^ and the post-acute sequelae of SARS-CoV-2 infection (PASC, defined as ongoing, relapsing, or new symptoms, or other health effects occurring four or more weeks after acute SARS-CoV-2 infection).^3^ In addition, serology testing has public health value and is used for epidemiologic purposes to assess the burden of prior SARS-CoV-2 infections in the population. However, the interpretation of antibody testing has been challenging for many reasons. First, assays use different technologies and measure different classes of immunoglobulins. Second, these assays are directed towards different SARS-CoV-2 proteins such as the nucleocapsid (N) or spike (S) proteins. While IgG anti-N antibodies reflect past infection irrespective of vaccination, the vaccines approved in pediatric populations induce the production of IgG anti-S antibodies. Thus, detection of these antibodies needs to be interpreted with caution in those with prior vaccination against COVID-19.

We conducted a retrospective study as part of the NIH Researching COVID to Enhance Recovery Initiative (RECOVER; https://recovercovid.org/), which seeks to understand, treat, and prevent PASC in children and adults. Leveraging PEDSnet, which is a multi-institutional clinical research network that analyzes EHR data from several of the nation’s largest children’s healthcare organizations, we sought to develop an accurate and reliable EHR-based algorithm to identify children and adolescents <21 years of who tested positive for SARS-CoV-2 infection exclusively by serology during the pandemic.^4^ In addition, we contrasted the serology-positive cohort with the PCR-positive cohort to identify differences in demographic and clinical characteristics. Identification of children that could have been paucisymptomatic and not tested or missed early in the pandemic is of critical importance to define the prevalence and burden of PASC in children.

## METHODS

### Data Source

Electronic Health Record (EHR) data was retrieved from all healthcare encounters at PEDSnet (pedsnet.org) institutions associated with children and adolescents who underwent serology and/or SARS-CoV-2 PCR testing in outpatient, inpatient, and emergency department (ED) settings. The institutions that participated in the study included Children’s Hospital of Philadelphia (CHOP), Cincinnati Children’s Hospital Medical Center, Children’s Hospital Colorado, Ann & Robert H. Lurie Children’s Hospital of Chicago, Nationwide Children’s Hospital in Columbus, Nemours Children’s Health System (a Delaware and Florida health system), Seattle Children’s Hospital, and Stanford Children’s Health.^5^ Institutional source data were standardized to the PEDSnet common data model, described in detail elsewhere^6^. CHOP’s Institutional Review Board designated this study as not human subjects’ research and waived informed consent.

### Cohort Formation

To define the serology-positive cohort, we first examined the frequency and type of serology testing performed at PEDSnet institutions for children and adolescents <21 years of age at the time of the health encounter, from March 1, 2020, to April 20, 2022. The serology tests included were IgM, IgG anti-N antibodies, IgG anti-S or receptor binding domain (RBD) antibodies, and IgG and IgA undifferentiated antibodies. We then identified children who tested positive for SARS-CoV-2 by serology only and did not have a positive SARS-CoV-2 PCR test. EHR documentation of COVID-19 vaccination has not been fully linked and harmonized with other EHR data within our network. Thus, to assure that serology tests results were related to a past SARS-CoV-2 infection rather than vaccination, we applied the timing of age-specific vaccine approvals by the US FDA and excluded children with positive IgG anti-S/RBD, IgA or undifferentiated IgG tests after the vaccination eligible periods. We used the following vaccine approval dates: December 12, 2020: BNT162b2 (Pfizer-BioNtech) in children > 16 years of age; May 12, 2021: BNT162b2 (Pfizer-BioNtech) for children 12-15 years of age, and November 2, 2021: BNT162b2 (Pfizer-BioNtech) for children 5-11 years of age. The adenovirus vaccine <u>(Ad26.COV2.S) marketed by</u> Janssen was approved in the U.S in February 28, 2021 for patients > 18 years of age. However, the overall uptake of this vaccine has been low, especially in the 18–25-year age group; thus, serology age-cut offs for this vaccine were not included. Cohort entry for the positive serology group was defined as the date of first positive IgM, IgA or IgG COVID-19 antibodies after applying the filters described above. For the PCR-positive group, cohort entry was defined as the date of a first positive SARS-CoV-2 PCR test, irrespective serology testing.^4,7^ COVID cases were defined as per the CDC guidance (https://services.cdc.gov/case-definitions/coronavirus-disease-2019-2021)

### Statistical analyses

We examined rates of monthly serology testing and percentage of positive serology tests over time using descriptive statistics and data visualization. We used chi-square tests to examine differences in demographic and clinical characteristics between the serology and PCR positive cohorts, and calculated effect sizes using standardized differences. Clinical characteristics of interest included chronic conditions, site of testing (ED, inpatient, outpatient) and COVID-19-related diagnoses. The Pediatric Medical Complexity Algorithm (PMCA) Version 2.0 was applied to categorize children as having no chronic conditions, non-complex chronic conditions, or complex chronic comorbidities, considering diagnoses up to 3 years before cohort entrance as described.^8,9^ We examined MIS-C or COVID-19 diagnosis recorded ± 30 days of the SARS-CoV-2 test. Analyses were conducted using R version 4.1.2 (2021-11-01).

## RESULTS

### Serology Testing Over Time

From March 1^st^, 2020, through April 20, 2022, we identified 18,647 serology tests and 1,764,658 PCR tests performed in 1,025,349 unique patients within the PEDSnet network. The same patient could have had more than one test performed in the data set. As such, we identified 348,678 patients that had multiple PCR tests performed, and 3,225 patients with multiple serology tests performed during the study period. The serology tests most commonly used were undifferentiated IgG antibodies (7,361 tests; 40%) and IgG anti-N antibodies (7,326 tests; 39.3%), followed by IgM antibodies (2,108 tests; 13.3%), anti-S or RBD IgG antibodies (1,127 tests; 6%), and undifferentiated IgA antibodies (637 tests; 3.4%).

The number and types of tests performed at each participant site are included in **Table S1**. The use of undifferentiated IgG or IgG anti-N antibodies was relatively high throughout the study, with a peak in testing during the winter of 2020-21 (**Fig 1A**). The use of IgM was highest in the spring of 2020 and decreased over time, whereas the use of IgG anti-S/RBD antibodies increased following vaccine approval for adolescents in December 2020. The overall percentage of positive serology tests increased over time and peaked during the omicron wave from December 2021 – February 2022 (**Fig 1B**).

**Figure 1.**
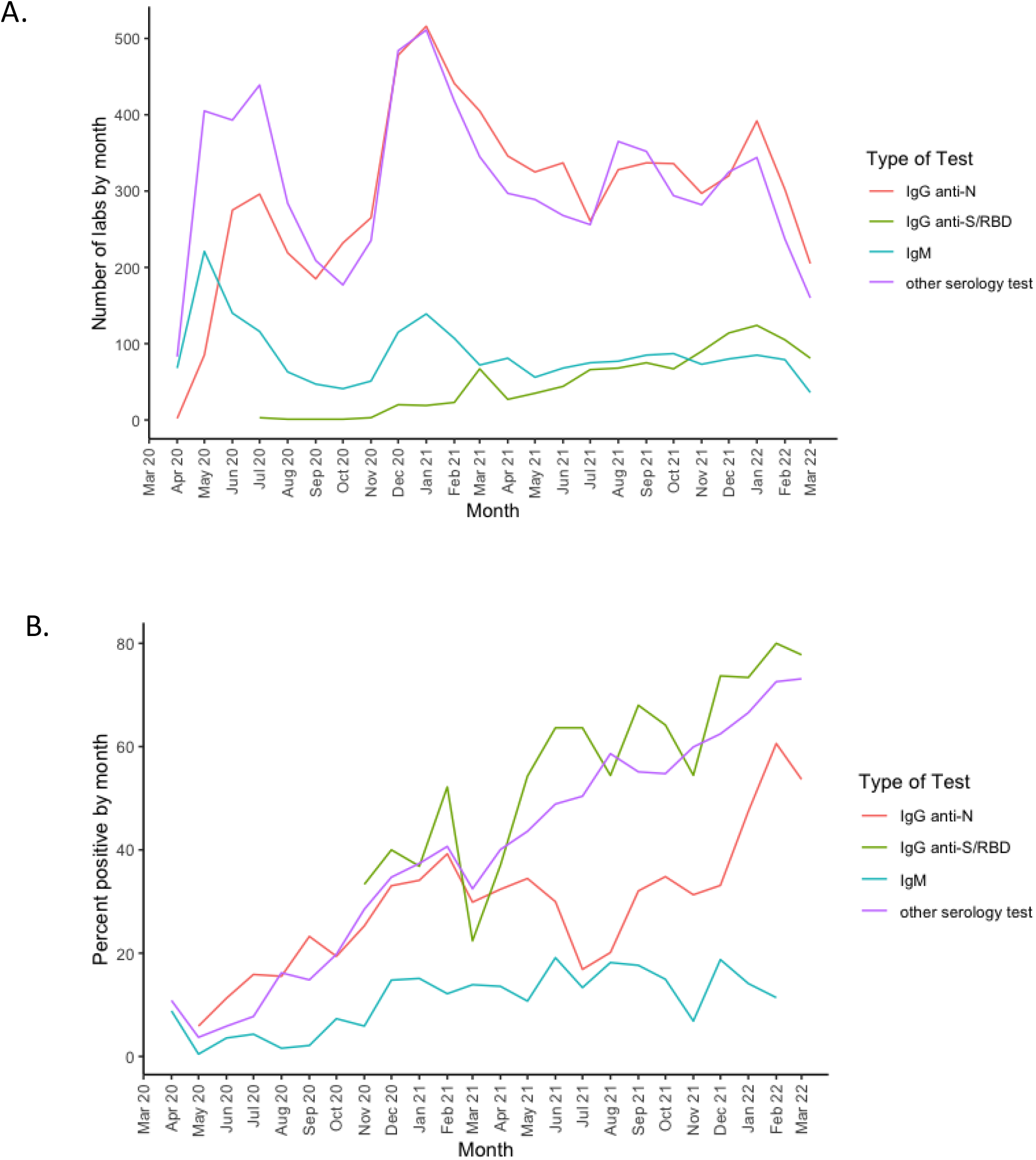
Number and positivity rates of different SARS-CoV-2 serology tests performed during the COVID-19 pandemic. **A**. Monthly number of serology tests performed with a valid (positive or negative) result. The Y axis represents the number of tests performed by type of tests over time (X axis). **B**. Monthly percentage of positive tests by type of test are plotted in the Y axis. IgG anti-N antibodies are depicted in red, IgM antibodies in blue, IgG anti-S/RBD antibodies in green and other serology tests, that included undifferentiated IgG and IgA antibodies, are depicted in purple.

Positivity rates for IgG anti-N or IgM antibodies reflected changes in SARS-CoV-2 infections, including known increases during the alpha variant phase (March-June 2021), the circulation of the delta variant (July-December 2021), and the circulation of omicron that began in December 2021 based on CDC COVID-19 data.^10^ Increases in the positivity rates of serology tests able to detect anti-S antibodies were particularly pronounced in children 12-15 and >16 years of age the months following age-specific approvals for COVID-19 vaccines (**Fig 2**).

**Figure 2.**
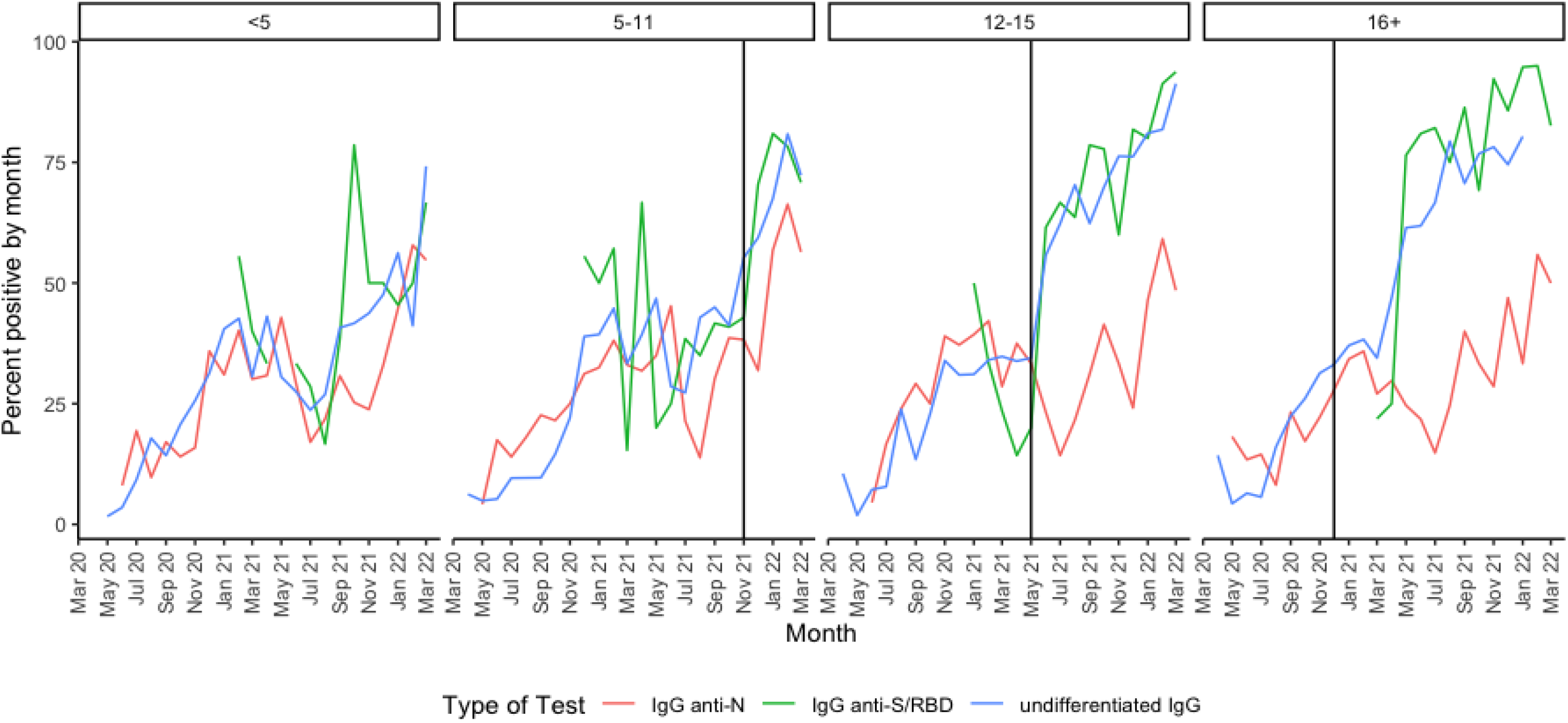
Positivity rates of SARS-CoV-2 antibody tests according to age. Percentage of serology tests with positive results by month (IgG anti-N in red, undifferentiated IgG shown in blue and IgG anti-S/RBD shown in green) are depicted according to age that was stratified in 4 groups: < 5 years, 5-11 years, 12-15 years and 16 to < 21 years of age. Vertical lines indicate the date of vaccine approval for each specific group.

### Contrast between the serology and PCR positive SARS-CoV-2 cohorts

Of the 1,025,349 patients with at least one PCR or serology test performed in serum or plasma specific for SARS-CoV-2, <0.1% of values were excluded because of unknown results or missing values. After excluding negative test results, there were 135,661 unique patients with at least one positive PCR, a positive serology test result, or positive for both tests at any time during the study period. Of those 131,537 (97%) were identified by PCR, and 4,124 (3%) exclusively by serology testing. There were 792 children that tested positive for COVID-19 by both PCR and serology during the study period. These children remained in the PCR cohort, as they would have been identified by our PCR-based algorithms. After applying age-specific vaccine cut-offs, we identified 2,714 patients who tested positive by serology and did not have a positive PCR test reported (**Fig 3**).

**Figure 3.**
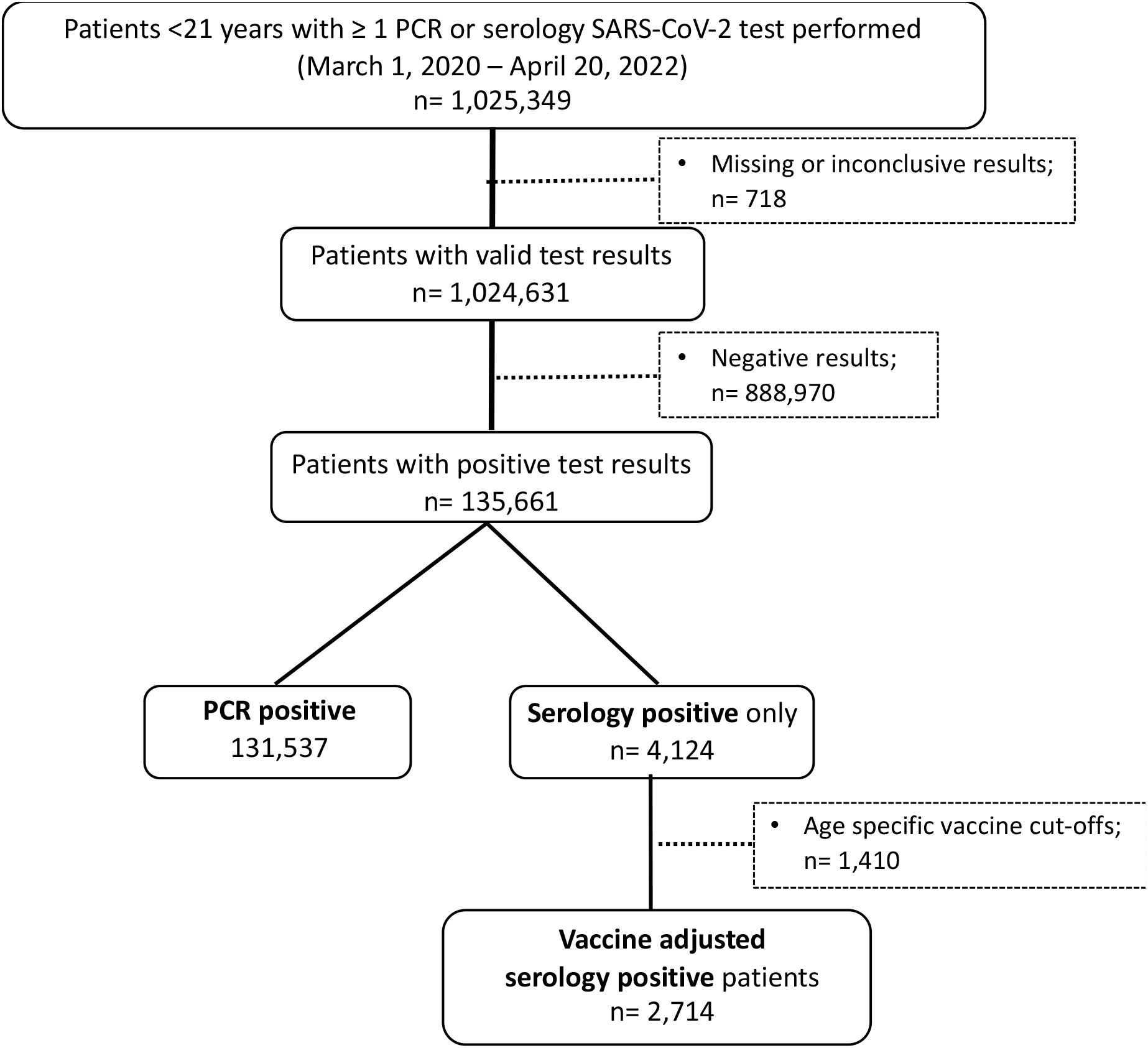
Flow diagram of patient selection based on serology and PCR testing. Of all patients with a COVID-19 test performed, after excluding missing data, negative or inconclusive results, there were a total of 135,661 patients with a positive test result (97% by PCR and 3% by serology testing exclusively). Of those, one third were excluded after applying age-specific cut-offs for vaccine approval.

The characteristics of the 1,024,631 children and adolescents that underwent testing for SARS-CoV-2 by PCR and/or serology, and of the 134,251 that tested positive are reported in **Table 1**. Compared to PCR-positive patients, serology-positive children were more likely to be Non-Hispanic White (55% vs. 45%), and a greater proportion were in the 12-15 years of age group (23% vs. 18%), largely reflecting the characteristics of those who were serology-tested (**Table S2**). Serology-positive patients were also more likely to have a chronic condition compared with PCR-positive patients (37% vs. 24%), especially complex medical conditions (27% vs. 11%). Most serology and PCR-positive patients were tested in the outpatient setting (52% and 76% respectively), but a higher proportion of serology-positive patients were tested as inpatients compared with PCR-positive patients (24% vs. 2%). Patients identified solely by serology as compared to PCR were more commonly diagnosed with MIS-C (23% vs <1%). On the other hand, a COVID-19 diagnosis was significantly more common in the PCR-positive than in the serology-positive cohort (72% vs. 45%).

**Table 1.** Demographic and Clinical Characteristics of the SARS-CoV-2 positive PCR and serology Cohorts

|  |  | **SARS-CoV-2**<br>**infection**<br>**n = 134,251** | **SARS-CoV-2**<br>**PCR cohort\***<br>**n = 131,537** | **SARS-CoV-2**<br>**serology cohort**<br>**n = 2,714** | **SD**<br>**(p-value)** |
| --- | --- | --- | --- | --- | --- |
| **Age at cohort entrance, mean (SD)** |  | 8.7 (6.0) | 8.7 (6.0) | 9.5 (5.6) |  |
| **Age at cohort entrance, n (%)** | 0-4 | 46722 (35%) | 45959 (35%) | 763 (28%) | 0.18 (<.001) |
|  | 5-11 | 43109 (32%) | 42179 (32%) | 930 (34%) |  |
|  | 12-15 | 23862 (18%) | 23234 (18%) | 628 (23%) |  |
|  | 16-20 | 20558 (15%) | 20165 (15%) | 393 (15%) |  |
| **Sex, n (%)** | Female | 65101 (48%) | 63836 (49%) | 1265 (47%) | 0.04 (.05) |
|  | Male | 69138 (52%) | 67689 (51%) | 1449 (53%) |  |
| **Race/ethnicity, n (%)** | Hispanic | 25230 (19%) | 24733 (19%) | 497 (18%) | 0.26 (<.001) |
|  | Non-Hispanic White | 60252 (45%) | 58746 (45%) | 1506 (55%) |  |
|  | Non-Hispanic Black/AA | 25900 (19%) | 25549 (19%) | 351 (13%) |  |
|  | Non-Hispanic Asian/Pacific Islander | 4534 (3%) | 4452 (3%) | 82 (3%) |  |
|  | Other/ Unknown | 12904 (10%) | 12739 (10%) | 165 (6%) |  |
|  | Multiple | 5431 (4%) | 5318 (4%) | 113 (4%) |  |
| **Health institution, n (%)** | A | 29009 (22%) | 28389 (22%) | 620 (23%) | 0.49 (<.001) |
|  | B | 28807 (22%) | 28577 (22%) | 230 (9%) |  |
|  | C | 21645 (16%) | 21280 (16%) | 365 (13%) |  |
|  | D | 6787 (5%) | 6545 (5%) | 242 (9%) |  |
|  | E | 23172 (17%) | 22668 (17%) | 504 (19%) |  |
|  | F | 15308 (11%) | 14907 (11%) | 401 (15%) |  |
|  | G | 3213 (2%) | 2983 (2%) | 230 (8%) |  |
|  | H | 6310 (5%) | 6188 (5%) | 122 (4%) |  |
| **Chronic conditions, n (%)** | None | 101899 (76%) | 100184 (76%) | 1715 (63%) | 0.44 (<.001) |
|  | Non-complex | 17627 (13%) | 17369 (13%) | 258 (10%) |  |
|  | Complex | 14725 (11%) | 13984 (11%) | 741 (27%) |  |
| **Period of cohort entrance, n (%)** | Mar 2020-Jun 2020 | 2999 (2%) | 2944 (2%) | 55 (2%) | 0.44 (<.001) |
|  | Jul 2020-Oct 2020 | 9029 (7%) | 8832 (7%) | 197 (7%) |  |
|  | Nov 2020-Feb 2021 | 31239 (23%) | 30505 (23%) | 734 (27%) |  |
|  | Mar 2021-Jun 2021 | 10526 (8%) | 10017 (8%) | 509 (19%) |  |
|  | Jul 2021-Dec 15, 2021 | 30489 (23%) | 29870 (23%) | 619 (23%) |  |
|  | Dec 16/2021-April 20/2022 | 49969 (37%) | 49369 (37%) | 600 (22%) |  |
| **Test location, n (%)** | ED | 28894 (21%) | 28244 (22%) | 650 (24%) | 0.72 (<.001) |
|  | Inpatient | 3389 (3%) | 2745 (2%) | 644 (24%) |  |
|  | Outpatient | 101952 (76%) | 100532 (76%) | 1420 (52%) |  |
| **Diagnosis, n (%)** | MIS-C | 877 (1%) | 253 (<1%) | 624 (23%) | 0.82 (<.001) |
|  | COVID-19 (no MIS-C) | 96466 (72%) | 95230 (72%) | 1236 (45%) |  |
|  | No COVID-19 or MIS-C | 36908 (27%) | 36054 (28%) | 854 (32%) |  |

SD: standardized difference was used to measure the effect size between the PCR and serology positive cohorts and may be interpreted as equivalent to a Z-score of a standard normal distribution. \*Patients included in the PCR cohort could have a serology test performed as well.

## DISCUSSION

In this study, we developed and applied an EHR-based codeset to identify children with COVID-19 based on serology testing who would have been missed otherwise if cohort selection was based solely on PCR.^4^ We found that the frequency of serology testing was significantly lower than PCR, and that there was substantial variability in both the frequency and type of serology tests used across eight large pediatric institutions. Through data harmonization and exclusion of results during age-based vaccine eligibility periods, we identified a subset of children who had not been identified by molecular testing in our network. These patients were frequently tested in the inpatient setting, and one fourth had an MIS-C diagnosis compared to <1% for the PCR positive cohort. In addition, these children had a higher prevalence of underlying complex medical conditions compared with those identified by PCR. Accurate identification of children and adolescents with positive SARS-CoV-2 serology results is critical in studies evaluating the presentation, burden and risk of PASC in pediatrics.

Unlike nucleic acid amplification tests which detect SARS-CoV-2 RNA, serologic assays measure the antibody response to current or past infection and to vaccines. Antibodies against SARS-CoV-2 are typically detected more than two weeks of symptom onset, which limit their diagnostic utility during the acute disease.^11^ On the other hand, they are useful clinically to identify patients with post-acute manifestations of COVID-19 such as MIS-C and/or other post-acute sequalae of SARS-CoV-2 infection. This is especially relevant in children whose initial infection may have not been detected because of an asymptomatic, or mildly ill presentation that did not prevent them from developing PASC.^12^ Compared to the latest national estimates of SARS-CoV-2 cumulative infection in children, that are approaching 75% of the pediatric population,^13^ serology testing for COVID-19 was performed in a minority of children in our network. Thus, it is clear that the number of SARS-CoV-2 infected children is significantly greater than those captured in healthcare systems, and the numbers identified in our study likely represent the tip of the iceberg.

In our study, the tests that were used more commonly were undifferentiated IgG, followed by IgG anti-N antibodies. However, we observed that IgG anti-S antibody use increased after vaccine implementation. The FDA does not recommend antibody testing to evaluate the level of immunity against SARS-CoV-2 at any time and especially after vaccine administration. Whether the increased use and positive rates of IgG anti-S antibodies reflect the performance of these tests to examine vaccine responses, or whether they represent a true increase in the rates of SARS-CoV-2 infections in unvaccinated children, warrants further studies.

The diagnosis of PASC relies on a broad range of new, recurring or persistent clinical manifestations that last four or more weeks after SARS-CoV-2 infection and might be difficult to recognize in children.^14^ This coupled with the underreporting and underestimation of COVID-19 in pediatrics has limited our ability to fully define the long-term impact of SARS-CoV-2 infection in children. Understanding the utility and implementation of serology testing in children is critical and will provide comprehensive and essential information to define the computable phenotype of PASC in children. Nevertheless, a homogeneous, robust and reliable system to report SARS-CoV-2 infections is needed and would allow defining the true incidence of PASC in children, with symptoms that might be underrecognized and/or attributed to other long-standing health conditions.

Our study has limitations. We lack the reasons that prompted caregivers to order serology testing which would require manual chart review. On the other hand, it is reassuring that one fourth of positive serology patients had an associated MIS-C diagnosis, suggesting that these tests were ordered in an appropriate clinical context. We took a conservative approach and used vaccine approval dates to exclude children with positive serology tests that did not allow us to differentiate between SARS-COV-2 infection and vaccination. Thus, we have likely underestimated and not included a group of children with true COVID-19. Future studies will incorporate vaccination registry data to overcome this limitation. A minority of children within our network underwent serology testing, and results suggest that it was biased to sicker children. Future studies should incorporate a combination of approaches, including PCR testing, serology testing, and diagnoses to be able to define the computable phenotype of PASC in children. Despite these limitations, we included a large sample size of children across eight major pediatric healthcare systems in the US, with variable practices, that allowed us to capture a pediatric population at risk for PASC.

In conclusion, data harmonization and development of a refined COVID-19 EHR-based serology system has enabled additional identification of children with prior COVID-19 infection, beyond PCR testing alone, that might be at risk for PASC. This work is necessary to capture relevant cohorts that will allow us to examine the presentation, risk, and burden of PASC in children.

## Data Availability

Data will be available upon reasonable request to the authors

## Abbreviations

COVID-19: coronavirus disease 2019
SARS-CoV-2: severe acute respiratory syndrome coronavirus 2
EHR: electronic health record
ED: emergency department
N: nucleocapsid
S: spike
RBD: receptor binding domain
PCR: polymerase chain reaction
PASC: post-acute sequelae of SARS-CoV-2 infection
MIS-C: multisystem inflammatory syndrome in children
FDA: Food and Drug Administration--FDA
CDC: Center for Diseases Control and Prevention

## Conflicts of Interest

AM reports funding from Janssen, Merck for research support, and Janssen, Merck and Sanofi-Pasteur for Advisory Board participation; SR reports prior grant support from GSK and Biofire and is a consultant for Sequiris. RJ is a consultant for AstraZeneca, Seqirus and Dynavax, and receives an editorial stipend from Elsevier.

## Role of funder/sponsor statement

The funder had no role in the design and conduct of the study; collection, management, analysis, and interpretation of the data; preparation, review, or approval of the manuscript; and decision to submit the manuscript for publication.

## Notes

### Competing Interest Statement

Asuncion Mejias reports funding from Janssen, Merck for research support, and Janssen, Merck and Sanofi-Pasteur for Advisory Board participation; SR reports prior grant support from GSK and Biofire and is a consultant for Sequiris. Ravi Jhaveri is a consultant for AstraZeneca, Seqirus and Dynavax, and receives an editorial stipend from Elsevier.

### Funding Statement

This research was funded by the National Institutes of Health (NIH) Agreement OT2HL161847-01 as part of the Researching COVID to Enhance Recovery (RECOVER) program of research.

### Author Declarations

Childrens Hospital of Philadelphia Institutional Review Board designated this study as not human subjects research and waived informed consent

