## Supplementary Table S1 and S2 for "Leveraging serology testing to identify children at risk for post-acute sequelae of SARS-CoV-2 infection: An EHR-based cohort study from the RECOVER program": Supp. Tables.docx

**Table S1. Number of serology tests performed at each PEDSnet site from March 1, 2020 to April 20,2022**

| **Site** | **IgG undifferentiated** | **IgG anti-N** | **IgM** | **IgG anti-S or RBD** | **IgA** | **Ab undifferentiated** |
| --- | --- | --- | --- | --- | --- | --- |
| A | 3,680 | 0 | 640 | 0 | 640 | 0 |
| B | 460 | 660 | 20 | 100 | <11 | 80 |
| C | <11 | 1,560 | 300 | 510 | 0 | <11 |
| D | 190 | 800 | 0 | <11 | 0 | 0 |
| E | 0 | 2,570 | 0 | 110 | 0 | 0 |
| F | 1,530 | 490 | 0 | 100 | 0 | 0 |
| G | 0 | 1,240 | 0 | 250 | 0 | 0 |
| H | 1,500 | 0 | 1,150 | 60 | 0 | <11 |
| **Total** | 7,361 | 7,326 | 2,108 | 1,127 | 637 | 88 |

Site-specific counts were rounded to the nearest 10 to mask counts of <11.

**Table S2. Characteristics of children and adolescents based on COVID-19 testing**

| Characteristic | | Patients with PCR tests performed*  n = 1,020,336 | Patients with serology tests only performed  n = 4,295 |
| --- | --- | --- | --- |
| Age at cohort entrance, mean (SD) | | 7.9 (5.8) | 11.9 (5.4) |
| Age at cohort entrance, n (%) | 0-4 | 404422 (40%) | 632 (15%) |
|  | 5-11 | 327592 (32%) | 1301 (30%) |
|  | 12-15 | 160547 (16%) | 1183 (28%) |
|  | 16-20 | 127775 (12%) | 1179 (27%) |
| Sex, n (%) | Female | 485534 (48%) | 2103 (49%) |
|  | Male | 534606 (52%) | 2192 (51%) |
| Race/ethnicity, n (%) | Hispanic | 165180 (16%) | 464 (11%) |
|  | Non-Hispanic White | 514902 (51%) | 2899 (68%) |
|  | Non-Hispanic Black/AA | 150348 (15%) | 171 (4%) |
|  | Non-Hispanic Asian/Pacific Islander | 43139 (4%) | 232 (5%) |
|  | Other/ Unknown | 103986 (10%) | 403 (9%) |
|  | Multiple | 42781 (4%) | 126 (3%) |
| Health institution, n (%) | A | 177602 (17%) | 774 (18%) |
|  | B | 205124 (20%) | 407 (9%) |
|  | C | 171967 (17%) | 343 (8%) |
|  | D | 73104 (7%) | 245 (6%) |
|  | E | 155404 (15%) | 672 (16%) |
|  | F | 122344 (12%) | 695 (16%) |
|  | G | 57139 (6%) | 683 (16%) |
|  | H | 57652 (6%) | 476 (11%) |
| Chronic conditions, n (%) | None | 777710 (76%) | 2800 (65%) |
|  | Non-complex | 127894 (13%) | 569 (13%) |
|  | Complex | 114732 (11%) | 926 (22%) |
| Period of cohort entrance, n (%) | Mar 2020-Jun 2020 | 72645 (7%) | 435 (10%) |
|  | Jul 2020-Oct 2020 | 199793 (20%) | 654 (15%) |
|  | Nov 2020-Feb 2021 | 204316 (20%) | 945 (22%) |
|  | Mar 2021-Jun 2021 | 152517 (15%) | 723 (17%) |
|  | Jul 2021-Dec 15, 2021 | 265237 (26%) | 989 (23%) |
|  | Dec 16/2021-April 20/2022 | 125828 (12%) | 549 (13%) |
| Test location, n (%) | ED | 209984 (21%) | 453 (11%) |
|  | Inpatient | 51291 (5%) | 187 (4%) |
|  | Outpatient | 758900 (74%) | 3655 (85%) |
| Diagnosis, n (%) | MIS-C | 915 (<0.1%) | 99 (2%) |
|  | COVID-19 (no MIS-C) | 438607 (43%) | 1295 (30%) |
|  | No COVID-19 or MIS-C | 580814 (57%) | 2901 (68%) |

*Patients included in the PCR cohort could have a serology test performed as well. COVID-19 diagnosis data includes codes associated with confirmed as well as suspected or presumptive COVID-19.
